# Digital Pathology, Deep Learning, and Cancer: A Narrative Review

**DOI:** 10.1101/2024.03.14.24304308

**Authors:** Darnell K. Adrian Williams, Gillian Graifman, Nowair Hussain, Maytal Amiel, Tran Priscilla, Arjun Reddy, Ali Haider, Bali Kumar Kavitesh, Austin Li, Leael Alishahian, Nichelle Perera, Corey Efros, Myoungmee Babu, Mathew Tharakan, Mill Etienne, Benson A. Babu

## Abstract

**Background and Objective:** Cancer is a leading cause of morbidity and mortality worldwide. The emergence of digital pathology and deep learning technologies signifies a transformative era in healthcare. These technologies can enhance cancer detection, streamline operations, and bolster patient care. A substantial gap exists between the development phase of deep learning models in controlled laboratory environments and their translations into clinical practice. This narrative review evaluates the current landscape of deep learning and digital pathology, analyzing the factors influencing model development and implementation into clinical practice.

**Methods:** We searched multiple databases, including Web of Science, Arxiv, MedRxiv, BioRxiv, Embase, PubMed, DBLP, Google Scholar, IEEE Xplore, and Cochrane, targeting articles on whole slide imaging and deep learning published from 2014 and 2023. Out of 776 articles identified based on inclusion criteria, we selected 36 papers for the analysis.

**Key Content and Findings:** Most articles in this review focus on the in-laboratory phase of deep learning model development, a critical stage in the deep learning lifecycle. Challenges arise during model development and their integration into clinical practice. Notably, lab performance metrics may not always match real-world clinical outcomes. As technology advances and regulations evolve, we expect more clinical trials to bridge this performance gap and validate deep learning models’ effectiveness in clinical care. High clinical accuracy is vital for informed decision-making throughout a patient’s cancer care.

**Conclusions:** Deep learning technology can enhance cancer detection, clinical workflows, and patient care. Challenges may arise during model development. The deep learning lifecycle involves data preprocessing, model development, and clinical implementation. Achieving health equity requires including diverse patient groups and eliminating bias during implementation. While model development is integral, most articles focus on the pre-deployment phase. Future longitudinal studies are crucial for validating models in real-world settings post-deployment. A collaborative approach among computational pathologists, technologists, industry, and healthcare providers is essential for driving adoption in clinical settings.

## 1. Introduction

Cancer remains one of the leading causes of morbidity and mortality worldwide (1,2). In 2019, the World Health Organization estimated that cancer is the leading cause of death globally. According to the National Center for Health Statistics, there will be 1,958,310 newly diagnosed cancer cases in 2023 (3). The American Cancer Society projects 609,820 cancer deaths in 2023 (3).

Screening and treatment have improved but remain a considerable primary public health concern. Public health systems worldwide spend $200 billion on cancer-related costs (4). By 2030, there will be 19.3 million new cancer cases and 10 million cancer deaths (5). As these numbers continue to rise, there is a need for enhanced efforts to diagnose and treat cancer for all patient populations. To ensure equitable access to care for all patient populations, addressing and overcoming disparities and biases in medical care is essential, thus providing patients with the care they deserve (6). Four principles underlie health equity: equal access, equal utilization of resources, health equality, and distribution according to need (7). Health disparities can be reduced by addressing multi-level structural determinants and ensuring access to disadvantaged populations (8). The Joint Commission and the Institute for Healthcare Improvement oversee initiatives and prioritize inclusion and fairness in clinical practice (9,10).

Cutting-edge healthcare innovations that transform glass slides into digitized formats, when coupled with artificial intelligence (AI) and telecommunication systems, have the potential to ensure equitable access to cancer care, particularly in regions facing shortages of specialists. Digital pathology (DP) originated in the late 1960s, with telepathology (TP), a branch of DP, as one of its first uses. TP involves sending digital images over a secure long-range network. 1986 Ronald Weinstein, MD, coined the term TP (11). Since then, validation studies have shown DP and conventional light microscopy to be highly accurate (11–15). Diagnosis does not require an onsite pathologist. TP has also eradicated the need for long-distance travel for specialized care, effectively addressing pathologist shortages and resource limitations within these networks. This innovation promotes equity in healthcare by making specialized care more accessible. TP also facilitates international expert consultations, enhances provider communication, and ensures seamless handoffs. Since the 1960s, the evolution of digital images has been remarkable, progressing from expensive devices to sophisticated robotic microscopes and autonomous robotic whole slide image (WSI) intelligent scanners with integrated storage and retrieval systems, significantly enhancing workflow efficiency (16). The appeal of digitizing slides with WSI technology lies in its convenience, portability, and the power to manipulate and analyze pixels. Furthermore, WSI systems are utilized for virtual education and novel research and serve as indispensable tools for primary cancer diagnosis (17).

In the era of precision medicine, integrating deep learning (DL) with these technologies is crucial for enhancing health equity and advancing the field. This can be achieved by developing DL models to identify algorithmic biases in cancer care (18). Furthermore, digital slides have been successfully utilized in pathology, leveraging computational pathology techniques to deliver faster and more precise outcomes. (19–21). DL algorithms in DP can help pathologists identify complex patterns, classify image features, and provide quantitative assessment and predictive analytics. By harnessing advanced digital image management systems using cloud-based technology, healthcare organizations can facilitate faster, quality workflows (16,17). As this technology progresses, the anticipated global revenue for digital pathology is projected to soar to a remarkable $2,045.9 million by 2029, boasting a robust Compound Annual Growth Rate of 12.6% (22). Amidst this rapid expansion, a significant opportunity emerges to enhance clinical outcomes in precision oncology. This article offers an overview of the literature surrounding digital pathology (DP) and deep learning (DL) models in oncology. The DL development lifecycle encompasses critical stages, including data preprocessing, model development, deployment, and continuous management within clinical practice. Additionally, we delve into the factors that influence model development. We present this article in accordance with the Narrative Review reporting checklist.

## 2. Methods

This study follows the PICO Framework. Problem: human samples for cancer diagnosis. Intervention: WSI and DL. Comparison (Evaluation): Model evaluation. It’s not possible to compare each model directly due to the differences in data sets, algorithms, and output metrics used in each study—outcomes: model performance metrics such as accuracy, F1 score, and others to measure model performance.

We searched Embase, PubMed, DBLP, Google Scholar, IEEE Xplore, Cochrane database, IEEE, Web of Science, ArXiv, BioRxiv, MedRxiv Web of Science, and Semantic Scholar.

Articles were published between 2014 and 2023, using terms 1) Boolean Logic: connecting words like “AND,” “OR,” and “NOT” in various combinations to expand or narrow down search results 2) Fuzzy Logic: search terms like “Digital Pathology” NEAR “Deep Learning” or “Whole Slide Imaging” WITHIN 5 words of “Deep Learning” to search for particular articles 3) Truncation: We searched for terms that began with a specific string by placing an asterisk (*) at the end of a root word. The word string used in the search were digital pathology, deep learning pathology, cancer, digital AI pathology, artificial intelligence, and cancer.

Studies on cancer detection utilizing WSI and DL were considered eligible Table 1. Employing the specified search criteria, a total of 776 articles were identified. Two independent reviewers, BB and AR, screened the articles to determine eligibility based on the inclusion criteria. Out of the initial pool, thirty-six papers were deemed eligible for inclusion in the study. In cases of disagreement, a third reviewer, BKK, served as a tiebreaker to reach a consensus. Figure 1. Shows the flow chart of studies using PRISMA (Preferred Reporting Items for Systematic Reviews and Meta-analysis) guidelines.

**Figure 1.**
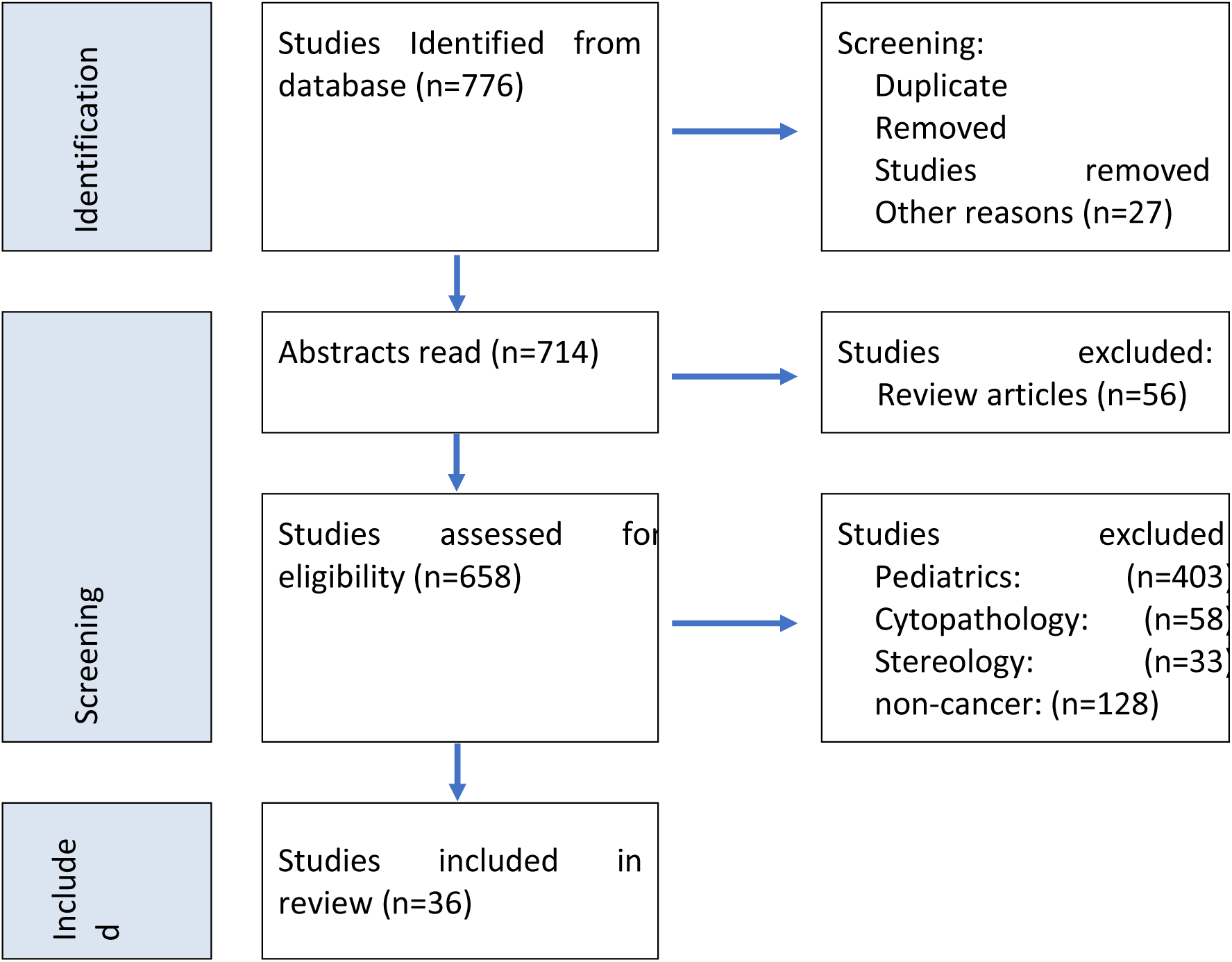
Flow chart of studies using PRISMA guidelines.

**Table 1.** The search strategy summary.

| Items | Specification |
| --- | --- |
| Date of Search | The first search was conducted on 9/18/2022. The last search was conducted on 5/18/2023. |
| Databases and other sources searched | The Databases searched were Embase, Pubmed, DBLP, Google Scholar, IEEE Xplore, Cochrane database, IEEE, Web of Science, ARxiv BioRxiv, MedRxiv Web of Science, and Semantic Scholar. |
| Search terms used | Search Using terms 1) Boolean Logic: connecting words like "AND," "OR," and "NOT" in various combinations to expand or narrow down search results 2) Fuzzy Logic: search terms like "Digital Pathology" NEAR "Deep Learning" or "Whole Slide Imaging" WITHIN 5 words of "Deep Learning" to search for particular articles 3) Truncation: We searched for terms that began with a specific string by placing an asterisk (*) at the end of a root word. The word string used in the search were digital pathology, deep learning pathology, cancer, digital AI pathology, artificial intelligence, and cancer |
| Timeframe | 2014-2023 |
| Inclusion and exclusion criteria | **Inclusion Criteria**<br>● Age greater than 18 years old<br>● Dates Range 2014 to 2023<br>● DL Novel Models<br>● Peer & Non-Peer Reviewed Publications<br>● Cancer<br>**Exclusion Criteria**<br>● Pediatric population<br>● Cytopathology<br>● Stereology<br>● Non-Cancer<br>● Review articles |
| Selection process | Using the search criteria, 776 articles were found. Two independent reviewers (BB) and (AR) selected articles that fit the inclusion criteria. Thirty-six papers met the study eligibility criteria out of 776. A third reviewer (BKK) served as a tiebreaker to resolve disagreements. |

## 3. Discussion

### 3.1 Model Types

The upcoming section offers a categoric overview of the DL models used in the literature. This will include exploring the various types of DL methods and analyzing their model development processes. By examining these aspects, we aim to provide a thorough understanding of the capabilities of these DL models in cancer detection. The selected literature is denoted in Table 2.

**Table 2.** lists the 36 articles. Dates ranged from 2014 to 2023.

| Study | Model | Performance Metrics | Training Data | Problem Type | Cancer Type | Staining Type |
| --- | --- | --- | --- | --- | --- | --- |
| Wang et al. (23) | Patch-Based with Various DL models | Two evaluation metrics: Lesion-based (based on correctly identifying cancer cells in WSI): 0.7051<br>Slide-based: 0.925 ROC | Camelyon-16 dataset: 400 | Classification | Breast | Pan cytokeratin |
| Wang et al. (24) | Patch-Based Paired with FCN-RF (Context-Aware Block Selection) Spatial Contextual Information) | M4-CNN Weighted Norm3-Based RF acc: 0.973 | SUCC dataset<br>Training: 355<br>Testing: 1402 | Classification | Lung | H&E |
| Chikontwe et al. (25) | MIL Patch Aggregation, Center Embedding | F1: 92.36<br>Precision: 92.54<br>Recall: 92.31<br>Accuracy: 92.31 | 173 WSI | Classification | Colon | H&E |
| Xie et al. (26) | MIL Patch-Based End-to-End (Learning Diverse Discriminative) | 0.986 Accuracy | BCC Dataset<br>Training: 6900<br>Valid: 1487<br>Test: 1575<br>Prostate Needle Biopsies Dataset<br>Training: 8521<br>Valid: 1827<br>Test: 1811 | Classification | Prostate | H&E |
| Li et al. (27) | 2 Stage Patch Based MIMS CNN Scale Invariant | 0.986 Accuracy | 339 3D OCT<br>Training: 67%<br>Test: 33% | Classification | Colorectal | H&E |
| Tsuneki and Kanavati (28) | Patch aggregation Weakly Supervise Efficient Net B1 | Best model TL-colon poorly ADC-2 (x20, 512)<br>Accuracy: TUR-P<br>Hospital A-B: 0.916<br>Hospital A: 0.969<br>Hospital B: 0.874<br>TCGA: 0.821<br>Hospital A-C: 0.844 | Training: Hospital A: 281<br>Hospital B: 739<br>Validation on: Hospital A and B: 20<br>Test: Hospitals A and B: 500 | Classification | Stomach, Colon, Lung, and Breast | H&E |
| Kanavati et al. (29) | Patch aggregation RNN/CNN<br>Efficient Net B1 | DCIS: Test set1: WS + RNN: ROC AUC: 0.937<br>IDC: Test set1: WS + RNN: ROC-AUC: 0.977<br>DCIS: Test set2: WS + RNN: ROC AUC: 0.960<br>IDC: Test set2: WS + RNN: ROC-AUC: 0.959 | Set: total WSI's: total Biopsy: total Surgery: Test: 1930, 1065, 865 Train: 1652, 978, 674 Valid: 90, 58, 32 | Classification | Breast | H&E |
| Campanella et al. (30) | MIL-RNN | AUC 0.98 | skin, prostate, and breast slides: 44,732 | Classification | Prostate, Basal Cell Carcinoma, and Breast | H&E |
| Shao et al. (31) | TransMIL | AUC 96.03% TCGA NSCLC<br>98.82% TCGA RCC | 993 NSCLC<br>884 RCC | Classification | Kidney, Breast, and Lung | H&E |
| Zhang et al. (32) | DTFDMIL | Camelyon-16 ACC 90.8%, F1 88.2%, AUC 94.6%<br>TCGA ACC 89.4%, F1 89.1%, AUC 96.1% | Camelyon-16 patches: 3.7 million<br>TCGA patches: 8.3 million | Classification | Lung | H&E |
| Wang et al. (33) | Second Order MIL | AUC 96% | 130 WSI | Classification | Breast | H&E |
| Hashimoto et al. (34) | DA-Domain Adversarial MIL | ACC 87%<br>Precision 97%<br>Recall 81% | Malignant Lymphoma Slides : 196 | Classification | Lymphoma | H&E |
| Lu et al. (35) | (CLAM) Cluster Constraint Attention Multiple Instance Learning | AUC: > 0.95 | Variable sizes used: 884, 1967, 899 | Classification | Lung | H&E |
| Sharman et al. (36) | Cluster-2-Conquer | C2C (w WSI+Patch+K LD Loss)<br>Accuracy: 86.2 | Gastro-intestinal Dataset 413 High-res Images.<br>Train: 65%<br>Valid: 15%<br>Test: 20% | Classification | Gastrointestinal | H&E |
| Guan et al. (37) | Node Aligned Graph-based MIL | Accuracy: 0.896<br>AUC ROC: 0.946 | Patches for NSCLC: 13904<br>Patches for RCC: 14116 | Classification | Lung and Renal | H&E |
| Xu et al. (38) | Graph-Based | Accuracy: 27.9, 0.78 Mean AUROC with 0.03 std | SIFT Flow Segmentation Dataset with 2688 Images and 33 Classes<br>TCGA BRCA 20% test and 80% Train and Val | Segmentation | Lung and Breast | H&E |
| Lu et al. (39) | Graph-Based | TCGA AUC: 75%<br>Two Independent Data Set AUC: 80% | TCGA Two Independent data sets<br>Training: 80%<br>Test: 20% | Classification | Breast | DAB |
| Zheng et al. (40) | Graph-Based | Best Models with Accuracy<br>CPTAC: 2-label: Resnet + GT: 0.935 +- 0.010<br>3-label: CL + GraphAtt: 0.835 +- 0.022<br>TCGA 2-label CL + GraphAtt: 0.911 +- 0.011<br>3-label CL + GraphAtt: 0.797 +- 0.026 | CPTAC: 2071<br>WSI's TCGA: 288<br>WSI's NLST: 665 | Classification | Lung | H&E |
| Lu et al. (41) | Attention Based MIL with Contrastive Learning Self Supervised Model | AUC ROC 0.968 ± 0.022 | H&E strained breast cancer with 400 images of size 2048x1536.<br>Train: 300 images<br>Valid: 100 images | Classification | Breast | H&E |
| Ilse et al. (42) | Attention Based MIL | Breast: AUC ROC 0.799<br>Colon: AUC ROC 0.968 | 58 Breast<br>100 Colon | Classification | Breast and Colon | H&E |
| Li et al. (43) | Dual-Stream MIL Self Supervised Contrastive Learning | Cameylon-16 ACC 86.82%, AUC 89.4%<br>TCGA ACC 91% AUC 96% | TCGA Cameylon-16 271 training 129 test | Classification | Breast and Lung | H&E |
| Khened et al. (44) | Ensemble | Dice score: 0.782 | Datasets with train<br>Camelyon-16: 270, 129<br>Camelyon-17: 500, 500<br>DigestPath: 660, 212<br>PAIP: 50, 40 | Segmentation | Breast, Colon, and Liver | H&E |
| Kalra et al. (45) | MEM Invariant Representation | TCGA ACC 84.84% | TCGA 2580 WSI | Classification | Lung Adenocarcinoma and Lung Squamous Cell Carcinoma | H&E |
| Chen et al. (46) | SISH Self Supervised MIL | SISH Macro Average: 45.51% | WSI: 22,385<br>Anatomic Sites: 13<br>Subtypes: 56 | Classification | 37 Cancers including Lung carcinoma | Variability According to Cancer |
| Niehues et al. (47)] | Self-Supervised Attention based MIL | attMIL AUROC: 0.94±0.02 | The dataset consists of 30 patients' data. The performance is then compared with previous studies. | Classification | Rectal and Colon | H&E |
| Yao et al. (48) | Unsupervised Siamese Network |  | 1500 Australian colorectal cancer patients<br>Training: 70%,<br>Valid: 10%,<br>Test: 10% | Classification | Lung | H&E |
| Muhammad et al. (49) | Unsupervised Deep Convolutional Autoencoder-Based Clustering Model | Autoencoder with combined MSE Loss and Reconstruction Clustering Error.<br>Best Cluster: 13 | 246 ICC slides | Clustering | Breast and Prostate | H&E |
| Zhu et al. (50) | Unsupervised K Means Clustering. | WSISA score: 0.703 (NLST), 0.638 (TCGA LUSC), 0.60 (TCGA GBM)<br>WSISA score: 0.703 (NLST), 0.638 (TCGA LUSC), 0.603 (TCGA GBM)<br>WSISA score: 0.440 (NLST), 0.397 (TCGA LUSC), 0.645 (TCGA GBM) | NLST Dataset: 404 Patients, 1104 WSI's.<br>TCGA Dataset is Divided into two parts.<br>TCGA LUSC: 121 Patients, 485 WSI's<br>TCGA GBM: 126 Patients, 255 WSI's<br>Training: 65%<br>Validation: 25%<br>Test: 20% | Clustering | Lung | H&E |
| Olsson et al. (51) | Conformal Prediction | With Conformal (0.1%) Error Detects: 85% Unreliable Predictions Without Conformal: 2% Error: 25% (Error in Atypical Prostate Tissue) | Prostate Slides: 7788<br>Training: 3059 | Classification | Entropy, Pseudo hyperplastic, Small-Cell Cancer | H&E |
| Tsuneki et al. (52) | Weakly Supervised Multi-organ Classification | Best Model x10EfficientNet B1: Avg Accuracy: 0.903 | Training 1k, 2k, and 4k WSI's<br>Validation on set is randomly chosen.<br>Test Dataset: WSI's: 4896 | Classification | Prostate | H&E |
| Mohammad et al. (53) | Weakly Supervised | CLAM<br>Acc: 87.04<br>AUC: 95.06 | iCAIRD Endometrial<br>Train : 1497<br>Valid: 499<br>Test: 911 | Classification | Endometrial | H&E |
| Aswolinsky et al. (54) | Neural Image Compression | TCGA AUC: 94%<br>TCIA AUC: 94%<br>Independent datasets AUC: 84%-98% | WSI: 2000 | Segmentation | Lung | H&E |
| Tellez et al. (55) | Neural Compression | Accuracy: 0.725 | Rectum: 74 WSI<br>Camelyon-16: 60 WSI<br>Tupac-16: 40<br>WSI Training: 50,000 (extracted) | Compression and Analysis | Breast | H&E |
| Sornapudi et al. (56) | Image Segmentation | CIN model Test Results<br>OU15/OU16 F1 : 93.5<br>ACC : 96.5<br>AUC : 95.5 | 3 Datasets with 50 WSI's each<br>OUI : 2998<br>True ROIs : 20,841<br>False ROIs OUI5 : 4915<br>True ROIs: 12,595<br>False ROIs OUI6: 4106<br>True ROIs :8601 | Segmentation | Cervical | H&E |
| Schmitt et al. (57) | CNN | ResNet 50 + CNN MBA: 50+ accuracy 56.1%<br>Slide Preparation Date 100% Slide Origin | Total : 42<br>Images : 7<br>Training : 80%<br>Test : 20% | Classification | Breast, Lung, Skin, and Gastrointestinal | H&E |

Legend: WSI: Whole Slide Image, ACC: Accuracy, AUC: Area Under the Curve, ROC: Receiver Operator Characteristic Curve, Valid: Validation ROI: Region of Interest, CNN: Convolutional Neural Network, CLAM, Cluster Attention Multiple Instance Learning, TCGA: The Cancer Genome, Atlas Program, TCIA: The Cancer Imaging Archive, CPTAC: Clinical Proteomic Tumor Analysis Consortium, TUPAC: Tumor Proliferation Assessment Challenge, CAMYELON16: Cancer Metastasis in Lymph Node Challenge 2016, H&E: Hematoxylin and Eosin, DAB Staining: 3'.3'-Diaminobenzidine NSCLC: Non-Small Cell Carcinoma, RCC: Renal Cell Carcinoma, CIN: Cervical Intraepithelial Neoplasia, ICC: Intrahepatic Cholangiocarcinoma

High-resolution slide images are gigapixels in size and require more memory and computational resources, so they cannot be incorporated directly into deep-learning models. Splitting high-resolution slide images into patches and developing patch aggregation strategies can help deep-learning models analyze large image datasets efficiently.

Slides can be divided into patches to train DL models (30). Models can be taught to recognize patch features and patterns. After training, models can predict labels for new patches or entire images and reconstruct the source image using patch aggregation strategies. Patch-based aggregation optimizes memory usage and DL efficiency.

### 3.2 Patch-based Aggregation Methods

Patch-based aggregation in machine learning is often used in image processing and computer vision tasks. It involves dividing an image into smaller, fixed-size pieces or “patches.” Each of these patches is then analyzed and processed independently (50,58). This approach allows for a more detailed and localized understanding of the image data; learning about each patch may capture unique features, whereas the patches themselves are part of the bigger picture and may or may not contain the region of interest for tumor analysis. Patch-based aggregation is beneficial in tasks like image classification, object detection, and texture analysis. For instance, histopathology imaging can help identify specific features in WSI, such as tumors or other abnormalities (29). The algorithm can make more accurate predictions or identifications by focusing on small image areas at a time. Moreover, this method can be combined with various machine learning models, such as convolutional neural networks. In such combinations, each patch is fed into the network, which learns to identify patterns or features within these smaller segments of the image. This can lead to more nuanced and detailed image analysis than processing the entire image (26,35).

### 3.3 Multiple Instance Learning

During training, bags are labeled in Multiple Instance Learning (MIL). Each bag has multiple instances, but only the bag label is known (25,26,30,35,36,57,59). A bag label is valid when the precise location of each instance is not essential, but the overall composition is. Bags are treated as single entities without explicitly modeling individual instances. For optimal performance, MIL requires large amounts of data; a minimum of 10,000 slides is recommended (30,57). MILs require many data points to capture patterns. Spatial relationships between instances are another MIL limitation.

Cancer classification using WSIs poses challenges because of their large pixel size and the annotation limitations when used on convolutional neural networks (26). Several techniques have been developed to overcome this limitation. An End-to-End, Part Learning-based approach can learn diverse and discriminative features to predict prostate and basal cell carcinomas (26). Furthermore, it defines multi-label lung cancer architectural subtypes for clinical decision support (26). Similarly, Cluster-to-Conquer overcomes the computational and algorithmic challenges posed by gigapixel-sized WSIs and the lack of MIL annotation (36). Convolutional neural networks (CNN) encoders and aggregation improve classification accuracy by learning slide-level label representations (36).

In small sample sizes and the large size of WSIs, the DTFD-MIL Double-Tier Feature Distillation MIL uses pseudo bags to virtually enlarge the number of bags and utilize a double-tier model to improve feature representation (32). This method outperforms other existing methods on CAMELYON-16 and TCGA lung cancer datasets (32).

Whole Slide Histopathology Images Survival Analysis WSISA is an aggregation method for predicting cancer survival in cases that are too computationally complex for traditional survival models (50). Through adaptive sampling, WSISA extracts hundreds of patches from each WSI. An aggregation method makes patient-level predictions based on cluster-level deep convolutional survival (50). The process is evaluated through experiments on different datasets related to Non-small-cell lung cancer and Glioma, demonstrating its ability to significantly improve prediction performance compared to existing state-of-the-art survival methods (50).

Using MIL approaches, an additional center-embedded aggregation technique accurately classifies colon cancer dataset images. This method learns both instance and bag-level embeddings by hierarchical pooling of features (25).

### 3.4 Spatially Aware MIL

A need for more spatial information limits MIL’s applications. Spatially aware MIL (saMIL) has been proposed to address this problem (59). CNN extracts features from instances within each bag. Combining these features creates a more accurate and interpretable model.

The dual-stream MIL with contrastive learning learns meaningful features from local and global pathology images (43). Two streams of the model are trained, one focusing on local patches and the other on the whole image, and a contrastive loss function is used to encourage the model to learn similar representations (43).

The multi-task MIL (MT-MIL) algorithm handles multiple related tasks simultaneously to capture complex relationships. MT-MIL assigns each task a different bag-level label, and the goal is to learn a model to predict these labels accurately, as used in the study looking at both the diagnosis and prognosis of early-stage invasive breast carcinoma (58).

Global and local features are extracted using a Multiscale Domain-adversarial Multiple instance CNN, automatically detecting tumor-specific features in a WSI (34). It addresses the difficulties associated with annotating tumor regions in WSIs, extracting global and local image features, and detecting image features against differences in staining conditions among hospitals/specimens for malignant lymphoma use cases (34).

Similarly, a framework for correlating MIL is (TransMIL), incorporating morphological and spatial information (31). The proposed method achieved faster convergence than state-of-the-art methods in various experiments (31). This framework improves weakly supervised tumor classification and cancer subtype identification (31).

A second-order learning model (SoMIL) with an attention mechanism and RNN learns the bag and extracts instance-level feature information trained on the breast cancer lymph node metastasis dataset (33).

### 3.5 Graph-Based

Using graph theory, graph-based methods model spatial relationships at the local and global levels to better define cells within the tissue structure and its functional relationships (38,39). Graph-based methods such as the SlideGraph model predict HER2 status in breast cancer and accurately identify and predict HER2-positive regions (39).

Node-aligned graph convolutional network representation and classification addresses the large gigapixel size of WSI (37). Prior approaches MIL combined with graph convolutional network (GCN), but non-ordered pooling may lose valuable information. Using a global-to-local clustering strategy, Node-Aligned GCN (NAGCN) builds correspondence across different WSIs, representing them with rich local structural information and global distribution (37). It performs better in cancer subtype classification datasets and can be applied to improve WSI representation (37).

The graph-transformer (GT) framework called GTP interprets morphological and spatial information for disease grade prediction (40). Contextual information is crucial in disease grading, overcoming the patch-based method’s limitations (40). GTP distinguishes adenocarcinoma and squamous cell carcinoma from normal histology (40). A graph-based saliency mapping technique called GraphCAM was also introduced, highlighting WSI regions associated with a class label (40).

### 3.6 Attention-Based MIL

An attention-based deep MIL for learning Bernoulli distributions of bag labels, where neural networks fully parameterize the bag label probability (42). Compared to other methods, the proposed method outperforms two real-life histopathology datasets without sacrificing interpretability (42). Pathologists highlight each instance’s contribution to each bag label to identify disease markers in large histopathological images (42).

Only sets are labeled on individual data instances in a permutation-invariant neural network called the Memory-based Exchangeable Model (MEM). Based on input sequences embedded in high-level features, the model learns interdependencies among instances using a self-attention mechanism (45). For the classification of two subtypes of lung cancer, the model achieved an accuracy of 84.84% by relying on toy datasets, point cloud classification, and lung WSIs (45). The MEM model can classify Histopathology images into different cancer subtypes (45).

The attention-based deep MIL also accurately classifies breast and colon cancer, and it also assists with cancer survival prediction, including high accuracy in predicting the survival of breast cancer patients (42,48).

### 3.7 Weakly Supervised Learning

Annotation by hand takes time, is tedious, and needs to be more scalable. MIL methods handle weakly labeled or unlabeled data by training on bags of instance (25,26,30,32,35,36,57). Classifying tumors with weakly supervised deep learning using patch-based models with regions of interest does not require manual annotation (24,35). Weakly labeled semantic segmentation is where only image tags are available as class annotations (19). Latent structured prediction encodes the presence and absence of classes and assigns semantic labels to superpixels (19). It shows improved accuracy in per-class classification compared to state-of-the-art methods (19).

Multi-class learning improves the model’s capability to classify complex structures. For weakly supervised image segmentation, representative structure cues in WSI identify glandular regions in endometrial cancer images (53).

### 3.8 Self-Supervised Learning

Studies using self-supervised learning overcome the labeling bottleneck. One study combines self-supervised feature learning using contrastive predictive coding (CPC) with regularized attention-based MIL (41). It achieves state-of-the-art performance for binary classification of breast cancer histology images with high accuracy and an area under the ROC curve reporting (41). Similarly, in dual-stream MIL networks, MIL-based methods effectively address WSI classification without localized annotations (41).

Its model accuracy can be improved with a novel MIL aggregator that models the relations of the instances in a dual-stream architecture with trainable distance measurement. Self-supervised contrastive learning was used to extract good representations for MIL, and a pyramidal fusion mechanism for multiscale WSI features was used to improve the accuracy of classification and localization (43).

Self-supervised contrastive learning can extract good representations for MIL and reduce cost bags’ prohibitive memory costs (43). The classification accuracy and localization are further improved through pyramidal fusion (43).

The study also uses SISH (self-supervised image search for histology) self-supervised deep learning to search WSI (46). The self-supervised model achieves constant search speed after being trained with only slide-level labels (46). The model encodes whole slide images into discrete latent representations. Furthermore, it leverages a tree data structure for fast searching followed by an uncertainty-based ranking algorithm for image retrieval. It identifies similar regions across multiple large diverse WSI datasets with strong performance accuracy (46).

Lastly, the study uses a self-supervised approach that highly accurately predicts biomarkers from WSI ross various institutions and scanner types (47). It utilizes self-supervised, attention-based multiple-instance learning (attMIL) to train models capable of identifying vital morphological characteristics within the WSIs. This methodology allows the model to concentrate on specific regions of the WSI and obtain insights from the data (47).

### 3.9 Unsupervised Learning

In unsupervised learning, models discover patterns from unlabeled data on their own. A study identified intrahepatic cholangiocarcinoma subtypes using unsupervised clustering (49). Deep convolutional autoencoders group tumor morphologies based on visual similarity (49). It identifies patterns in cancer tissue images without knowing the type of tumor (49). Tumor stroma is crucial in tumor growth, angiogenesis, and metastasis.; it was used to stratify ductal carcinoma in situ (49). These models predict overall survival (49).

### 3.10 Transfer Learning

Transfer learning in digital pathology involves using pre-trained CNNs trained on large-scale image data sets and then fine-tuning them on pathology image datasets. Transfer learning aims to overcome the limitations of pathology slide annotations and, by doing so, may improve performance and save time and computational costs (60–64). At times, performance declines when using specific pre-trained models, for example, ones trained on ImageNet, because it may incorporate the significant differences in the natural and the pathology image datasets, resulting in performance inefficiencies (65).

Prostate carcinoma is detected from TransUretral Resection of the Prostate (TURP) images using DL trained on large WSI datasets, then fine-tuned on smaller datasets. Using transfer and weakly supervised learning, they trained DL models to classify TURP WSIs into prostate adenocarcinoma and benign lesions. With these results, these DL models are suitable for diagnostic workflows (52).

The DL breast ductal carcinoma in situ classification model trains a CNN on a large, annotated image dataset and uses transfer learning to improve performance (66). It reports improved accuracy compared to traditional machine learning (66).

### 3.11 Ensemble Learning

Ensemble learning combines DL models to improve classification performance (44). Ensemble methods reduce the risk of over-fitting and improve overall model accuracy by training multiple models with different architectures or input data (44). One study’s system’s predictions may assist clinicians in making medical decisions and managing treatment (44).

### 3.12 Other Algorithms and Techniques

#### 3.12.1 NN192 Machine Learning Algorithm

Acs et al. (67) is a significant study in digital pathology, focusing on developing an automated NN192 algorithm for assessing tumor-infiltrating lymphocytes (TILs) in melanoma. The study addresses the challenges of standardization and subjectivity in TIL assessment by utilizing hematoxylin-eosin-stained sections. The automated scoring system, validated through a retrospective analysis of 641 melanoma patients, demonstrates that higher TIL scores correlate with better disease-specific overall survival. This highlights the potential of the automated TIL scoring system as an independent prognostic marker in melanoma. Furthermore, the study showcases the transformative power of digital technologies in pathology, improving the accuracy and efficiency of pathological assessments. This research contributes to melanoma prognosis and sets a precedent for the broader application of digital pathology in cancer care and research (67).

Aung et al. (68) examine how digital pathology, specifically in melanoma, can improve cancer diagnosis and prognosis. Using the NN192 machine learning algorithm, the study achieves an objective and precise assessment of tumor-infiltrating lymphocytes (TILs) compared to traditional visually based evaluations. The analysis of TILs in melanoma samples highlights the potential of digital pathology to provide consistent and reproducible results. This advancement is crucial in personalized medicine, where accurate biomarker assessment is essential for tailoring cancer treatment. The study emphasizes integrating digital technologies into pathology for more precise diagnoses and better patient outcomes in oncology.

#### 3.12 2 Neural Image Compression

Neural Image Compression (NIC) reduces gigapixel WSI images to extremely compact representations for training CNNs to predict image labels (54,55,69,70). Three encoding tools transform low-level vector embeddings of high-resolution WSIs: contrastive learning, reconstructional error minimization, and Bidirectional Generative Adversarial Network (BiGAN). BiGANs outperformed the other two unsupervised encoding mechanisms, with a Spearman correlation of 0.521 (55). NIC builds convolutional neural networks for gigapixel image analysis based on weak image-level labels (55). The first step is compressing gigapixel images using an unsupervised neural network, retaining high-level information, and suppressing pixel noise (55). CNN is trained on these compressed image representations to predict image-level labels, avoiding manual annotations (55). Two public histopathology datasets were evaluated with NIC and found to integrate global and local visual information while attending to areas of the input gigapixel images that overlap with human annotations (55). NIC classifies non-small cell lung cancer subtypes with high accuracy (54). In addition, compression algorithms can preserve important image features and accuracy (70).

#### 3.12.3 Conformal Prediction

Conformal prediction is a mathematical framework to assess the reliability of prediction systems for diagnosing and grading prostate biopsies. A model is trained to estimate its accuracy, and conformal prediction intervals are generated to quantify its uncertainty (68). Conformal prediction may improve the reliability and interpretability of AI-assisted pathology diagnosis (68). Conformal prediction led to a lower rate of incorrect cancer diagnoses and flagged a higher percentage of unreliable predictions than AI systems without conformal prediction (68). Conformal prediction can help augment medical providers’ decision-making in the clinical setting (51).

### 3.13 Elements Impacting Model Development

Factors that should be considered arise when developing a DL model (71–75). Awareness of these factors is crucial to ensure a successful development process.

### 3.14 Reducing GPU Use and Memory Consumption with Sustainable AI Technique

A DL model for WSI often needs a lot of computing power, especially when working with large-pixel data. GPU usage and memory consumption can be high, leading to longer training times and higher costs. DL techniques have been developed to address this issue, limiting GPU use and memory consumption. Compressed models, transfer learning, data augmentation, and batch normalization have emerged as effective techniques that reduce GPU usage and memory consumption while improving model accuracy, training times, and energy costs, reducing the carbon footprint (76,77). Preparation variability slide prep, staining, tissue preparation protocols, and scanners can affect DL performance. This involves processing enormous amounts of data with these variations, labeling ground truth, comprehensive image pre-processing, denoising, and WSI normalization. Normalization and data augmentation also adjust for non-cancer cells, necrosis, and inflammation. MIL, attention, and graph-based methods capture spatial relationships between these cells. Furthermore, malignant cells can be challenging to identify and classify due to non-cancerous cells and tissue artifacts; this is addressed by handcrafted feature extraction.

### 3.16 Data Augmentation

Data augmentation involves rotating, flipping, and scaling existing data to generate new training data. As a result, DL models perform well and require less training data, and it helps prevent overfitting and reduce memory and GPU utilization.

### 3.17 Defining Accurate Ground Truth

We need ground truth and a labeled set of data points to train, test, and validate DL models to compare model outputs. The accuracy of ground truth impacts model accuracy. Experts traditionally label ground truth. Pathologists manually highlight regions of interest in images and train DL models to detect similar areas in other images. Expert annotation verification for large datasets is time-consuming, expensive, and not scalable. Experts may also need help labeling consistently.

### 3.18 Labeling Annotation

Labeling slide annotation is time-consuming and requires much effort from experts. Moreover, the amount of labeled data available may be limited, impacting the performance of DL algorithms. Recent progress in developing weakly, semi, self-supervised, and unsupervised machine-learning clustering techniques can be used to analyze WSI data without labeled data (24–26,30,32,35,36,41,43,46,47,49,53,57). In semi-supervised learning, a limited amount of labeled data is combined with a more significant amount of unlabeled data to train models; it allows DL models to perform better with less labeled data required.

### 3.19 Domain Adaptation and Diversity

Domain adaptation refers to the ability of a model to perform well on data from a target domain that may differ from the domain in which the model was initially trained. Poor domain adaptation is a significant challenge in WSI DL implementation, attributed to the high-resolution large gigapixel size of the images and annotation challenges. DL models may lose performance accuracy (35,78–80).

Data diversity, sensors, and patients must be considered when developing DL models for WSI. Managing data diversity is challenging since slide images can come from different hospitals, laboratories, and clinics. These differences in quality, resolution, and staining can affect DL models.

It is crucial to consider sensor type when training DL models. Sensors can also affect image quality. Images acquired by fluorescence microscopy may differ from those acquired by brightfield microscopy.

Finally, consider the patients; different patient populations must be represented in a diverse dataset.

### 3.20 Structured Prediction Global vs. Local Features

Structured predictions capture spatial and heterogeneous cell relationships in various spatial scales. Global cell features describe the overall distribution of cells in a tissue, while local cell features describe their immediate neighbors. This model captures complex spatial relationships between cells by combining these features. Interactions between cells are complex and dynamic (81). The properties of cells vary depending on where they are in tissues. By combining global and local features and examining their cellular relationships, structured prediction makes accurate predictions about individual cells’ malignant behavior.

### 3.21 High Spatial Resolution

A substantial amount of spatial data generated by WSI can make the development of accurate DL algorithms challenging. Studies reviewed noted that image compression and patch-based techniques can overcome this limitation.

### 3.22 Compressed Models

Model compression minimizes the number of parameters (54,55,69,70). A pruning technique removes less meaningful connections between nodes, and a quantization technique reduces the precision of the weights and activations. Compressed models use less memory, can be trained faster, reduce GPU usage, and improve digital storage (54,55,69,70).

### 3.23 Batch Normalization

During training, batch normalization and activations at each neural network layer are normalized to improve stability and convergence. Using batch normalization, DL models can be trained faster and more accurately and use less memory.

### 3.24 Transfer Learning

A pre-trained model is fine-tuned in transfer learning for a specific task (60–64,82). This approach can significantly reduce training time, GPU, and memory use. Transfer learning is especially effective for tasks with similar features and datasets. Recent developments in resource-efficient neuroevolutionary multi-tasking have shown promising results within machine learning pipelines, including AutoML (76). Gradient-free evolutionary optimizers have emerged as a powerful alternative to traditional DL, as demonstrated by OpenAI (83).

Combining ResNet50 (residual network 50), a convolutional neural network that is 50 layers deep, with weakly supervised or unsupervised techniques in DL WSI improves image classification performance and addresses gradient explosion. Training with a pre-trained ResNet50’s backbone is faster and more efficient. The model has already learned general features from a large dataset, requiring fewer data and iterations to reach good accuracy. This is useful for small datasets or scenarios requiring real-time inference. The quality and diversity of the training data determine the model’s performance. Data preparation and selection are critical to achieving optimal results. Not all pre-trained models are suitable for pathology. For example, DL models pre-trained on ImageNet at certain times may not be ideal for the computational pathology task (65). The ImageNet models are trained on natural images, whereas pathological images have unique features. To achieve optimal performance, pre-trained models may need to be fine-tuned or trained from scratch. Furthermore, this model may not capture all the variations in pathology images. Artifacts, stains, and other factors may affect the accuracy of DL models in pathology images. Careful evaluation of pre-trained models on pathology images and considering alternatives is essential.

### 3.25 Rare Conditions

Rare conditions are underrepresented in training data, affecting DL models’ ability to diagnose them. Techniques in transfer learning, data augmentation, synthetic data, and federated learning address these limitations.

### 3.26 Healthcare Bias, Inclusion, Fairness, and Equity

Deep learning digital services should incorporate inclusion, fairness, bias detection algorithms, metrics, and strict governing protocols during the DL model development lifecycle (6,18,84,85). DL model development workflows should strive to obtain model parity. Fletcher describes three aspects of DL model development: fairness, appropriateness, and bias (85). To limit disparities, these elements must be addressed when developing DL models: religion, economic status, ethnicity, race, and gender should also be incorporated into these algorithms (84). If left unaddressed, AI models may demonstrate bias, as noted by (84). Feedback algorithms can rectify skewed patient datasets (84). Local governance committees and agencies, such as the Joint Commission and Institute for Healthcare Improvement, provide ongoing oversight to ensure equitable conditions. Continuous monitoring of quality data underscores the importance of equity in clinical practice.

### 3.27 False Positives Management

DL models for WSI improve sensitivity to detect cancer. However, with this improvement, false positive detection increases. While the models are better at detecting cancer, they may also flag areas as cancerous when they are not. Training the models on more extensive and diverse datasets may mitigate the risk of false positives and improve their accuracy. A study revealed that techniques can reduce and improve false positives (86). When interpreting the results of these models, it is vital to consider false positive detections.

### 3.28 Data Privacy

Privacy and security concerns arise with WSI and DL technologies. As WSI deals with large amounts of sensitive patient information, strong data privacy policies, encryption, and access control are indispensable. Differential privacy methods such as encryption and federated learning protect individual privacy while allowing accurate data analysis (87–89).

## 4. Conclusions

Data pre-processing and model development represent only a small fraction of the expansive DL lifecycle. A truly seamless journey from model development to clinical production environment is ideal. To achieve this seamlessly on a healthcare enterprise scale within digital pathology, a robust AI technical infrastructure, expert personnel, and a governance system for continuous oversight are needed.

The digitalization of pathology has trailed behind its radiology counterpart, which transitioned from analog films to digital systems decades ago. This lag is evident in the field of DL within digital pathology compared to radiology, as demonstrated by the numerous algorithm approvals by the FDA. While the technical intricacies of model development are vital, as discussed in this review, they are but one piece of the puzzle.

Future longitudinal clinical trials during the post-deployment production phase of the DL lifecycle are essential. These studies will offer invaluable insights into the real-world clinical impact of DL on cancer care, guiding us toward a future where innovative technology and healthcare intersect seamlessly to benefit patients.

Being a narrative review, this study design may exhibit more bias and is not intended to be as comprehensive as other methods.

## Data Availability

All data produced in the present study are available upon reasonable request to the authors

## Acknowledgments

None

## Footnote

Reporting Checklist: The authors have completed the Narrative Review reporting checklist.

Peer Review File:

Conflicts of Interest: All authors have completed the ICMJE uniform disclosure form. The authors have no conflicts of interest to declare.

Ethics Statement

The authors are accountable for all aspects of the work in ensuring that questions related to the accuracy or integrity of any part are appropriately investigated and resolved.

